# The Effect of Sex on median effective concentration of ropivacaine for ultrasound-assisted caudal block in the elderly undergoing Anorectal Surgery

**DOI:** 10.64898/2026.06.23.26356307

**Authors:** Fu Wang, Mingyu Qu, Wei Zhao, Yin He, Hongyan Zhou, Liang Zhang

## Abstract

**BACKGROUND:** Caudal block is widely employed in pediatric lower abdominal surgeries and adult anorectal surgical by its simplicity of operation, reliable efficacy, and high safety. Previous studies have found that the dose of ropivacaine for Caudal block in adults exhibits gender differences. However, it remains unclear whether such differences in the median effective concentration (EC50) of ropivacaine also exist in the elderly population.

**METHODS:** This is a double-blind, prospective study, We enrolled patients aged 60–80 years with ASA physical status I-Ⅱ who were scheduled for anorectal surgery under caudal anesthesia, and allocated them to 2 study groups according to their gender. Each participant received a single injection of 20mL ropivacaine. Using Dixon’s up-and-down sequential allocation, the initial concentration of ropivacaine was set at 0.35% and the subsequent concentrations were determined by the analgesic response of the previous patients to the pinprick testing. The concentration change was 0.025%. The EC50 of ropivacaine in each group was determined using the the up-and-down method and probit regression. The primary outcome was the EC50 (95% confidence interval [CI]) of the 2 groups. Data on the surgical time, analgesic duration, and adverse events during surgery were also recorded.

**RESULTS:** This study included a total of 40 elderly patients (20 male and 20 female). The EC50 of ropivacaine for caudal block in elderly male patients was 0.263% (95% CI: 0.179%–0.311%), while that in elderly female patients was 0.281% (95% CI: 0.161%–0.353%). The EC50 of ropivacaine for caudal block in elderly female patients was approximately 6.4% higher than that in elderly male patients.

**CONCLUSIONS:** There is a significant gender difference in the EC50 of ropivacaine for caudal block among the elderly, with elderly female requiring a higher EC50 than male.

**Trial registration:** The Chinese Clinical Trial Registry ChiCTR2200064111.

## Introduction

Caudal block is a kind of anesthesia in which local anesthesia is injected into the sacral epidural space to block the sacral nerve and is particularly well-suited for anorectal and perineal surgeries^[1]^. Compared with other anesthetic methods, caudal block offers distinct advantages including simplified operation, less traumatic, reliable efficacy, optimal postoperative analgesia, and high patient satisfaction^[2]^. However, due to the relatively large volume of the caudal canal, a substantial dose of local anesthetic is typically required to achieve adequate blockade. This inevitably increases the risk of local anesthetic toxicity, particularly in elderly patients. Age-related physiological changes, including alterations in sacral canal anatomy and modified responses to local anesthetics, may predispose this population to complications such as extensive sensory blockade and hemodynamic instability during caudal anesthesia^[3–4]^. Presently, the administration of caudal anesthesia in geriatric patients relies heavily on empirical clinical judgment due to the paucity of robust data on optimal ropivacaine dosing for this demographic. Precise local anesthetic dosing is critical, as it directly determines anesthetic effectiveness, perioperative hemodynamic profiles, and overall patient prognosis. Therefore, investigating the median effective concentration (EC50) of local anesthetics for caudal block is valuable—not only to reduce complications associated with local anesthetics but also to meet the current demands of precision anesthesia.

The EC50 of local anesthetics serves as an objective and quantitative measure for evaluating drug potency, providing critical guidance for clinical application. However, this parameter is significantly influenced by multiple factors, such as the physicochemical properties of the drug, the patient-specific variables and detection methodology, etc. Over the past two decades, increasing attention has been paid to sex-related differences in anesthesiology. Studies have reported that commonly used anesthetic agents—such as propofol, volatile anesthetics, neuromuscular blocking agents, and opioids—exhibit significant variations between sexes^[5–7]^. In contrast, research on the relationship between sex differences and local anesthetics remains scarce. While a few studies suggest potential differences in ropivacaines EC50 between sexes—such as elevated requirements in females during caudal blockade—these findings are primarily derived from middle-aged cohorts^[8]^. Given the physiological distinctions between aging and younger adults (e.g., reduced muscle mass, altered drug metabolism), extrapolating existing data to elderly populations is unreliable. To date, no randomized controlled trial has systematically investigated whether sex influences ropivacaines EC50 in elderly patients receiving caudal anesthesia.

Therefore, this study enrolled elderly patients undergoing hemorrhoid surgery under caudal anesthesia and employed Dixon’s up-and-down sequential method and probit regression analysis to investigate the influence of gender differences on the EC50 of ropivacaine for caudal blockade in elderly patients. The findings aim to provide evidence-based guidance for local anesthetic dosing in elderly patients receiving caudal anesthesia.

## 1. Materials and Methods

### 1.1 Ethics Statement

This prospective, single-center, randomized controlled trial was conducted in the Department of Anesthesiology from September 28th, 2022 to April 8th, 2024, following the principles of the Declaration of Helsinki (2013 revision). Approved by the ethics committee of Chongqing Traditional Chinese Medicine Hospital (Approval No.: 2021-KY-65) and registered in the Chinese Clinical Trial Registry (https://www.chictr.org.cn/showproj.html?proj=172746) (Registration No.: ChiCTR2200064111) in September 26, 2022. Written informed consent was obtained from all participants, and patient confidentiality was strictly maintained throughout the study.

### 1.2 Patient Enrollment

The patients aged 60–80 years with American Society of Anaesthesiologists physical status I or II who undergoing elective hemorrhoidectomy with caudal block were included. Exclusion criteria: (1) Severe hemorrhagic disorders; (2) Infection at the puncture site; (3) History of central nervous system surgery or diseases, including schizophrenia, epilepsy, Parkinson’s disease, or myasthenia gravis; (4) Local anesthetic allergy; (5) Diabetes mellitus; (6) Repeat anal surgery; (7) Hypertension, coronary artery disease, or cardiac conduction abnormalities, including sick sinus syndrome, atrioventricular block, and severe bradycardia (HR <50 bpm); (8) Body mass index (BMI) >30 kg/m². Termination criteria included: (1) Failure to complete the trial; (2) Caudal block failure, including unilateral block, accidental intravascular injection, or local anesthetic systemic toxicity (LAST); (3) Voluntary withdrawal during the study.

### 1.3 Grouping and Trial Protocol

All eligible patients were stratified by sex into elderly male (Group C-m) and elderly female (Group C-f) cohorts. Using the Dixon and Massey “up-and-down” sequential allocation method to determine the EC50 of ropivacaine for each group. Each participant received a single injection of 20 mL ropivacaine, the initial concentration of ropivacaine was set at 0.35% and the subsequent concentrations were determined by the analgesic response of the previous patients to the pinprick testing (with concentration increments or decrements of 0.025%).

### 2.4 Ultrasound-assisted Caudal Block

The patient was routinely monitored upon arrival in the preparing room, including electrocardiography, non-invasive blood pressure, pulse oximetry. Ropivacaine (75 mg / 10 ml Disorat®; Guangdong Jiabo Pharmaceutical Co., Ltd.) used for the caudal block was diluted with 0.9% saline to 20ml according to target concentration. All patients were placed in the left lateral decubitus position for caudal block administration. Initial anatomical landmark identification was performed to determine the approximate position (located at the equilateral triangle formed by the sacral cornu and bilateral superior lateral sacral crests). Subsequently, the ultrasonography was employed for the exact position (Figure 2). After identifying the puncture point, a 22G needle was advanced through the sacrococcygeal ligament into the caudal epidural space. Upon confirming loss of resistance and verifying absence of blood or cerebrospinal fluid upon aspiration, 20 mL of ropivacaine was administered slowly. The detailed technical approach was consistent with our previously published methodology^[9]^. We assessed the analgesic effectiveness of the caudal block using pinprick testing of the perineal anal area at 20 min after the caudal block. The anesthetic effects of the patients were classified as follows:

1. The caudal block was considered effective if there was no pain in response to the pinprick testing of the perineal anal area.
2. The caudal block was considered ineffective if the patient experienced numbness but still perceived pain in response to the pinprick testing of the perineal anal area.
3. The caudal block was considered a technical failure in cases of vascular puncture, local anesthetic toxicity, unilateral block, or no anesthetic effect of the caudal block in the patients.

### 2.5 Anesthesia management

This study employed a restrictive fluid strategy, with total intraoperative fluid administration typically limited to ≤1 L in the absence of significant bleeding or hemodynamic instability, to reduce the incidence of postoperative urinary retention.

For ineffective block or technical failure, the patients received rescue anesthesia, which included supplemental opioids, local infiltration anesthesia by the surgeon, or another caudal block. Perioperative adverse events were managed according to standardized protocols: intravenous atropine for bradycardia (heart rate <50 bpm), intravenous ephedrine and fluid infusion for hypotension (systolic blood pressure <90 mmHg or >30% decrease from baseline), ensuring patient safety throughout the perioperative period.

### 2.6 Determination of the EC50

The Dixon and Massey up-and-down sequential allocation method was been used to determine the EC50 of ropivacaine in each group. Based on our previous findings, we selected 0.350% ropivacaine as the initial concentration^[9]^. Subsequent patients received concentrations determined by the preceding patient’s response in the same group: the concentration was decreased by 0.025% if the caudal block was “effective”, or increased by 0.025% if “ineffective”. Cases of technical failure were excluded from analysis, with the subsequent patient repeating the same concentration. When the response transitioned from “ineffective” to “effective”, these two adjacent concentrations formed an independent pair. This continued until at least six valid pairs were obtained per group.

### 2.7 Primary/Secondary Endpoints and Quality Assurance

The primary endpoint was the EC50 of 20 mL ropivacaine for ultrasound-assisted caudal block in elderly patients with sex stratification. The secondary endpoints included the EC95 of ropivacaine for each group, duration of analgesia, and incidence of adverse events.

To ensure consistency, all blocks were performed by one experienced anesthetist using the same ultrasound probe (Philips, Lumify L12-4, United States).The ropivacaine concentration for caudal blocks was prepared by a nurse not involved in subsequent research procedures. Efficacy assessments were performed by the operating surgeons. Both study participants and outcome assessors remained blinded to the local anesthetic concentrations and group allocations throughout the trial.

### 2.8 Sample size calculation

This study employed the up-and-down sequential allocation method, which precludes a priori sample size calculation^[10]^. The trial termination criterion was achievement of at least six crossover pairs. Based on prior studies, 20-30 participants are required to estimate the EC50^[11–12]^. Consequently, the number of enrolled cases was not predetermined in advance.

### 2.9 Statistical analysis

Statistical analyses were performed using SPSS version 26 (IBM Corp., Armonk, NY, USA). Demographic characteristics and various intraoperative indicators were collected. Normally distributed continuous variables are presented as mean ± standard deviation (SD) and analyzed by independent samples t-tests, while non-normally distributed data are expressed as median with interquartile range (IQR) and analyzed by Mann-Whitney U tests. Categorical variables were analyzed using chi-square (χ²) tests. Probit regression analysis was employed to estimate the EC50, EC95 and corresponding 95% confidence intervals (CIs) of ropivacaine for each group. All tests were bilateral tests, with P<0.05 considered statistically significant.

## 2. Results

During the study period (September 2022-July 2023), 44 consecutive elderly patients fulfilling the predefined eligibility criteria underwent standardized ultrasound-assisted caudal block. Per protocol, four technical failures (three unilateral blocks, one complete sensory blockade failure) were excluded, yielding 40 analyzable cases (20 per sex stratum) for final efficacy assessment. (Figure 1).

**Figure 1.**
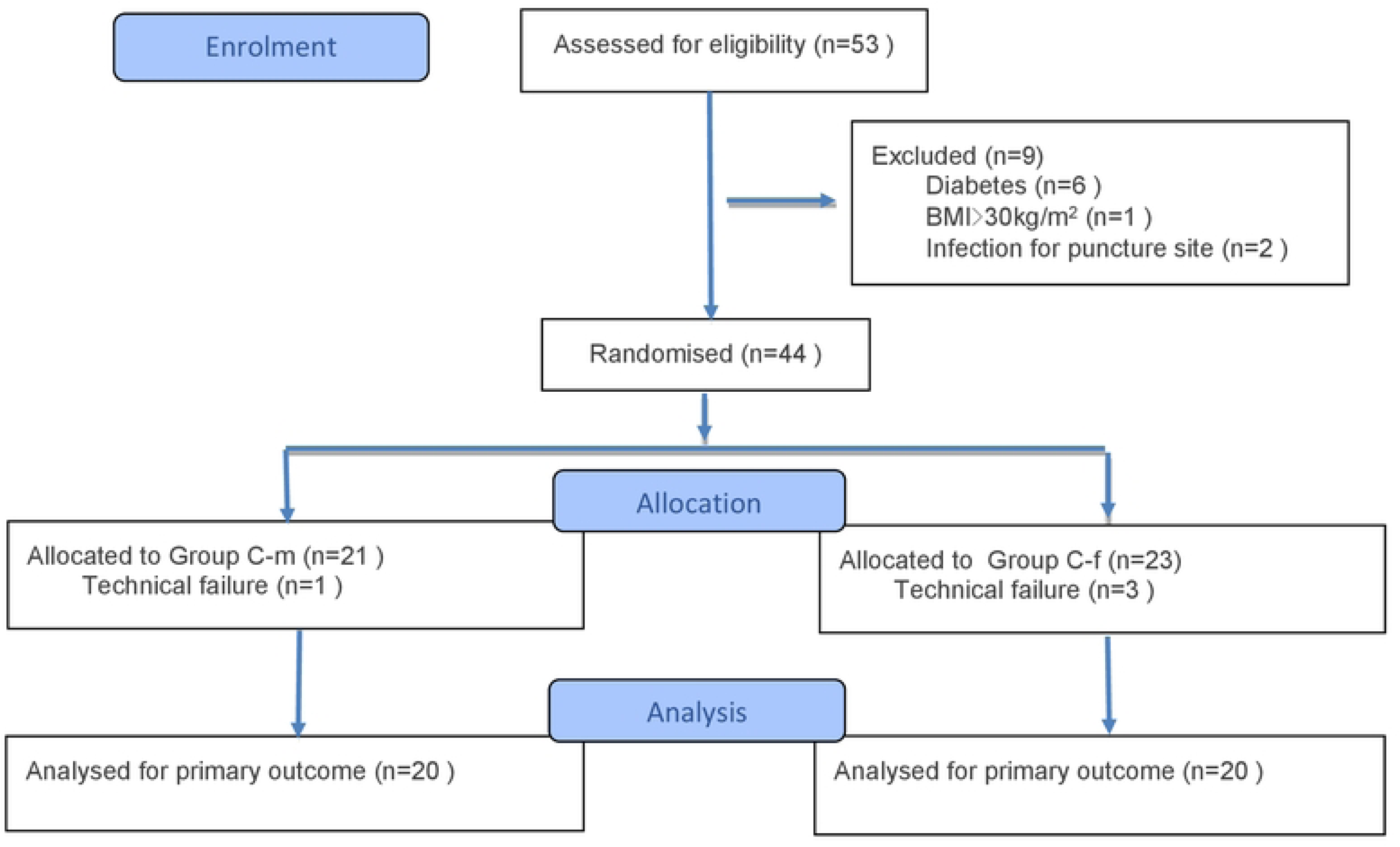
CONSORT 2025 Flow Diagram.

**Figure 2.**
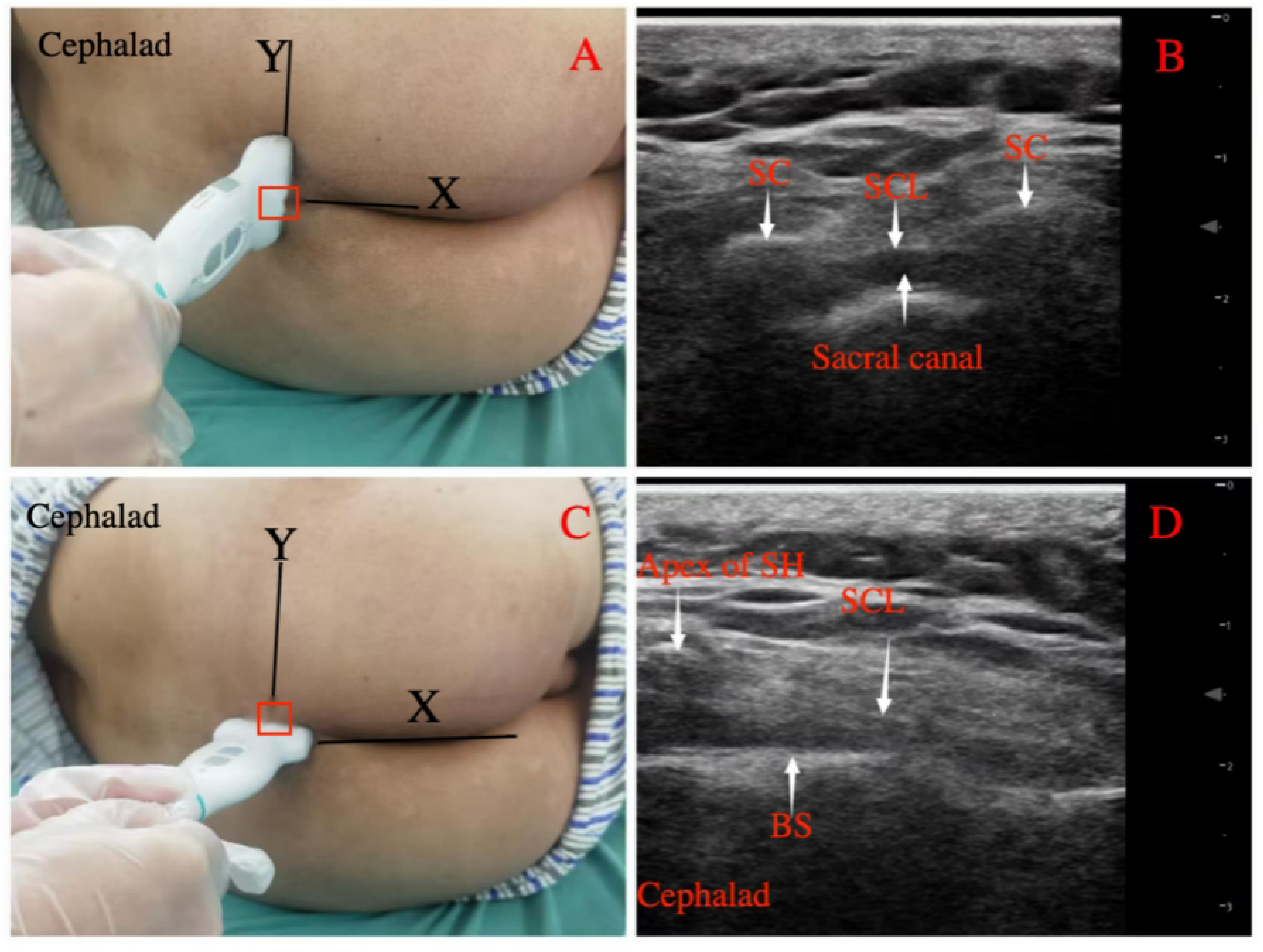
The Position of ultrasound transducer and The view.

### 3.1 Demographic data and clinical characteristics

The groups were well-matched in baseline characteristics, with comparable distributions of age (65.7 ± 3.7 years vs. 66.7 ± 5.9 years, P = 0.547), BMI (23.30 ± 1.89 vs. 24.15 ± 3.70, P = 0.367), intraoperative blood loss (12 ± 4.1 ml vs. 10 ± 2.8 ml, P = 0.08), surgical duration (45.8 ± 19.4 min vs. 42.4 ± 20.4 min, P = 0.587), and analgesia duration (520.5 ± 196.6 min vs. 557.1 ± 137.2 min, P = 0.498). (Table 1).

**TABLE 1.**
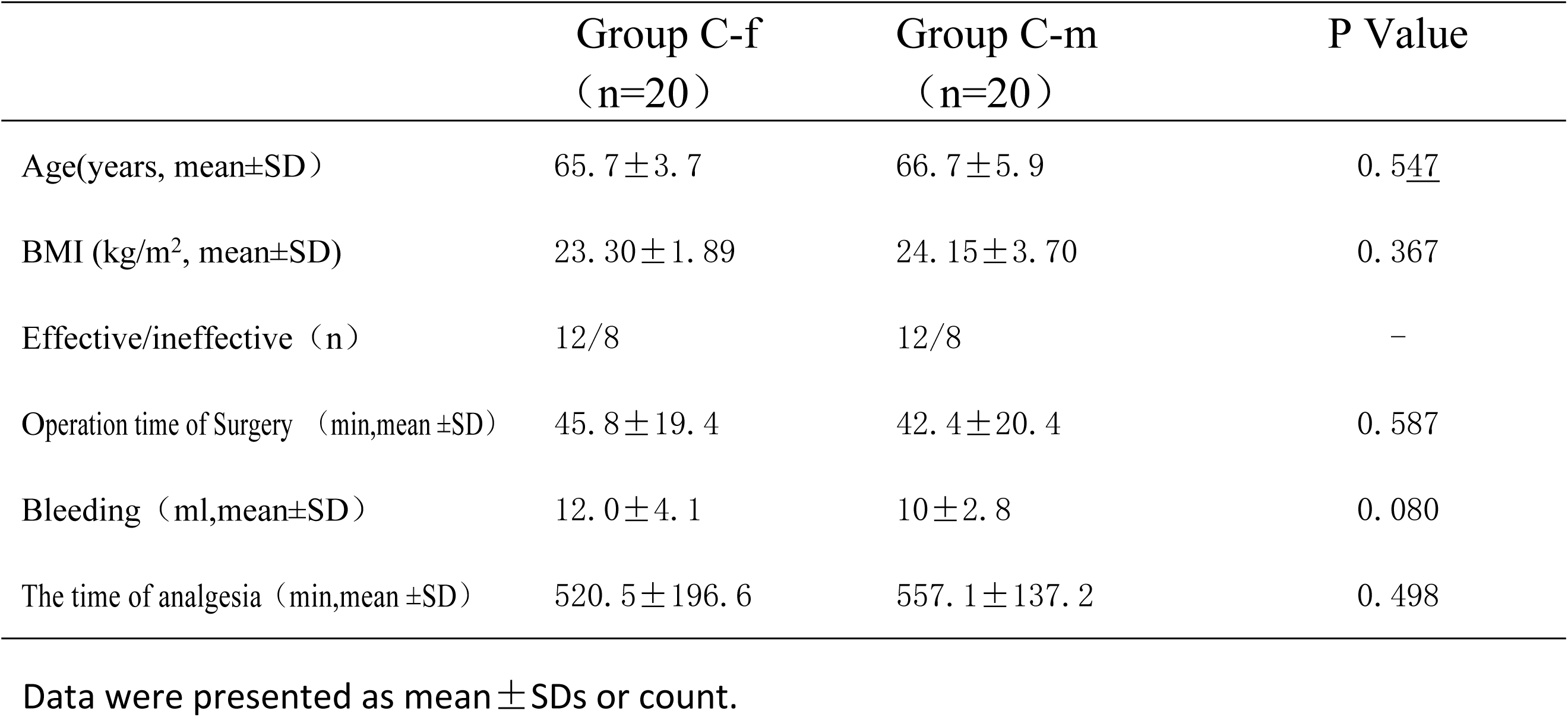
Demographic data and operation time.

### 3.2 EC50 and EC95 of Ropivacaine in the two Groups

Figure 3. displays the sequence of successful and failed outcomes using the up-and-down sequential allocation method. Probit regression-derived EC50 values (95% CIs) were significantly higher in Group C-f (0.281% [0.161–0.353%]) versus Group C-m (0.263% [0.179–0.311%]; The EC95 values of ropivacaine were 0.362% ( 0.282–0.562%) in group C-f and 0.338% (0.288–0.543%) in group C-m. (Table 2)

**Figure 3.**
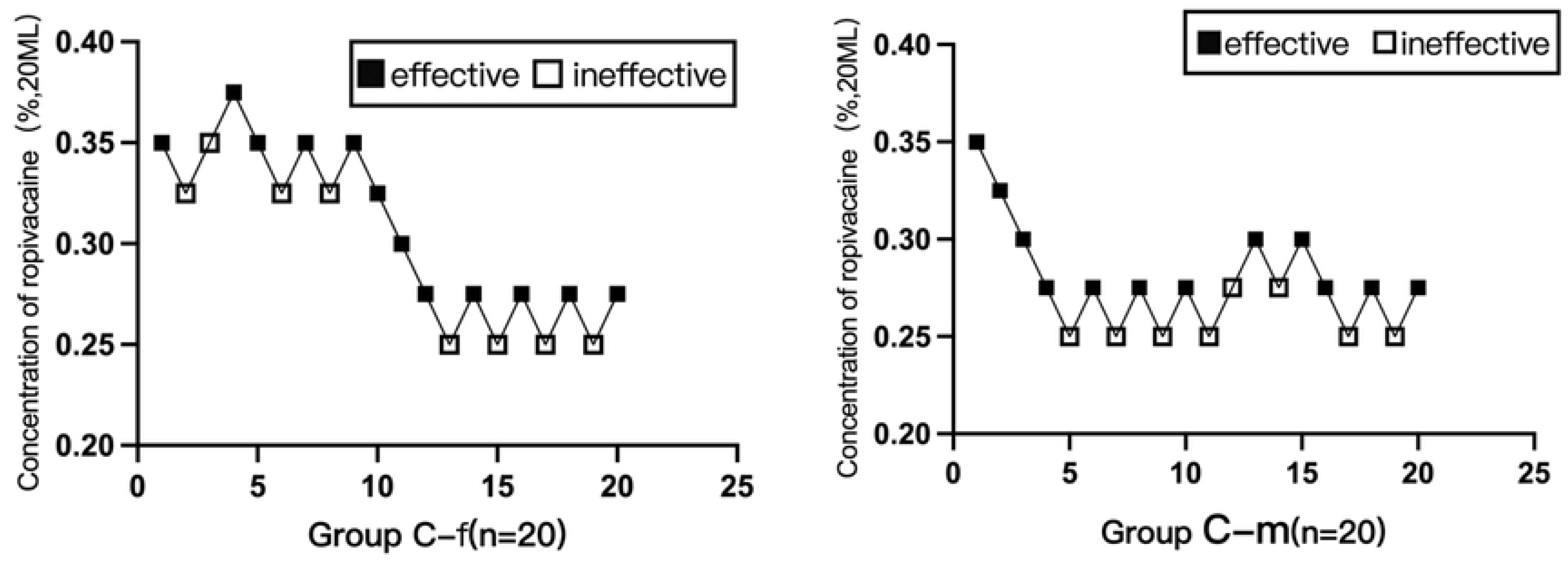
Dose-response concentrations of ropivacine for causes.

**TABLE 2.** Dose-response concentration of ropivacaine for caudal block derived by the probit regression.

| PROBIT Regression | Group C-f (%n=20) | Group C-m (%n=20) | P Value |
| --- | --- | --- | --- |
| EC<sub>50</sub><br>( and 95%CI) | 0.281<br>(95%CI=0.161~0.353) | 0.263<br>(95%CI=0.179~0.311) | 0.003 |
| EC<sub>95</sub><br>( and 95%CI) | 0.362<br>(95%CI=0.282~0.562) | 0.338<br>(95%CI=0.288~0.543) | 0.003 |

### 3.3 Adverse Reactions

No cases of drug-requiring bradycardia or severe hypotension were observed in either group.

## 4. Discussion

This represents the first study to systematically evaluate gender differences in ropivacaine requirements for caudal anesthesia in elderly patients. This study established that for primary hemorrhoid surgery under ultrasound-assisted caudal blockade with a fixed 20 mL ropivacaine volume, the EC50 was 0.263% in elderly male patients versus 0.281% in elderly female patients, demonstrating a statistically significant 6.4% higher requirement in female patients. These findings provide evidence-based reference for the medication of caudal block in elderly patients.

Gender has been confirmed as an important factor influencing the dosage of anesthetic drugs, recent studies have consistently demonstrated significant gender differences in ropivacaine sensitivity during caudal blockade. The research by Li et al. revealed that gender substantially influences the EC50 of ropivacaine for caudal anesthesia, with adult males requiring an EC50 of 0.296% compared to 0.389% in females - indicating markedly higher requirements in female patients^[7]^. Our current findings are consistent with these observations. Gender differences may stem from multiple factors. On one hand, physiological variations exist between males and females, including muscle mass, fat distribution, and vascular anatomy, which may influence drug distribution and diffusion^[13]^. On the other hand, sex hormone levels differ significantly between genders, potentially modulating drug effects through regulation of neurotransmitter systems and receptor expression^[14]^. Moreover, both qualitative and quantitative studies consistently conclude that females demonstrate higher sensitivity to various pain modalities and consequently require greater anesthetic doses compared to males^[15]^. However, in contrast to Li’s findings, the gender disparity observed in elderly patients was significantly less pronounced than in younger populations. This attenuation may be attributable to age-related changes in sex hormone levels. With aging, women experience a gradual decline in estrogen levels, and estrogen has been shown to reduce the pain threshold via the TRPV1 and ANO1 pathways^[16]^. Existing research corroborates that the magnitude of gender differences diminishes with advancing age^[17]^. Collectively, the observed variations in local anesthetic responses between sexes stem from a multifactorial interplay encompassing biological determinants, psychological influences, and sociocultural components.

Elderly patients often present with multiple comorbidities and demonstrate diminished physiological reserve and immune competence, resulting in significantly reduced tolerance to both anesthesia and surgical procedures. This study revealed that the median effective concentration (EC50) of ropivacaine for caudal blockade in elderly patients was significantly lower than in younger adults. This phenomenon may be attributed to age-related changes in sacral canal anatomy and altered responses to local anesthetics. First, with advancing age, the vertebral canal becomes more stenotic, and the permeability of the dura mater increases, facilitating enhanced diffusion of local anesthetics into the subdural space^[18]^. Consequently, even low doses may induce extensive neural blockade. Second, elderly patients frequently present with comorbidities that impair drug clearance while concurrently exhibiting reduced expression of voltage-gated sodium channels, rendering them more sensitive to local anesthetics^[19]^. Thus, comparable blockade can often be achieved with lower doses. Finally, progressive age-related degenerative changes in the nervous system—including reduced nerve fiber density—lead to elevated thresholds for pain and temperature perception^[20]^. Studies indicate that beyond 60 years of age, pain perception intensity declines by 10% to 20% per decade^[21]^. Furthermore, Li et al.’s study on lumbar epidural blockade demonstrated that advanced age significantly affects the EC50 of ropivacaine for motor blockade (0.383% in the elderly group vs 0.536% in the non-elderly group)^[22]^. Although this investigation focused on epidural rather than caudal blockade, the caudal approach remains a form of epidural anesthesia, thereby providing indirect evidence for age-related effects on caudal blockade. In summary, age constitutes a critical determinant of the effective local anesthetic concentration for ropivacaine in caudal anesthesia. Clinical practice should incorporate careful consideration of patient age, physiological status, and surgical requirements when selecting appropriate drug concentrations to optimize both anesthetic efficacy and patient safety.

It is well recognized that a successful nerve block depends on both an adequate volume of local anesthetic and precise needle placement. Therefore, this study adopts ultrasound-guided caudal block, which significantly improves positioning accuracy and procedural safety compared to traditional landmark-based techniques. This method is now widely used in both adult and pediatric caudal block procedures^[23]^. Jaffar et al. demonstrated that the success rate of sacral canal puncture based on anatomical positioning was approximately 70% to 75%, whereas Zhang et al. reported a 90% success rate with ultrasound-guided techniques, which is consistent with our study results^[24–25]^. Furthermore, the major complications of adult sacral canal block include: local anesthetic toxicity (with an incidence rate of approximately 0.92%) and sacrococcygeal puncture site pain (with an incidence rate of about 3.08%)^[26]^. Both complications are strongly associated with multiple puncture attempts during the procedure. Ultrasound guidance enables direct visualization of sacral anatomy, allowing for accurate identification of anatomical variations (structural anomalies, sacral cysts), precise needle placement into the target space, and avoidance of blind puncture-related complications (e.g., total spinal anesthesia)^[27–28]^. The significantly improved procedural precision substantially reduces unnecessary tissue trauma, leading to markedly diminished pain and discomfort during anesthesia^[27]^. Consequently, this study employed ultrasound-guided caudal blockade, which not only dramatically enhances the success rate of the procedure but also provides patients with a safer and more comfortable anesthetic experience.

## 5. Limitations of this study

This study had some limitations. Firstly, this study focused on patients aged 60–80 years with mixed hemorrhoids, as our primary objective was to investigate gender differences in ropivacaine requirements for caudal blockade in the elderly population. The exclusion of patients >80 years was primarily due to safety concerns. As demonstrated in prior research, super-aged patients (≥80 years) exhibit increased susceptibility to local anesthetics due to factors such as higher comorbidity burden, frailty, sarcopenia, and reduced hepatic/renal function, which collectively lower the threshold for local anesthetic systemic toxicity (LAST) and elevate perioperative risks compared to younger elderly individuals^[29]^. Secondly, our study defined successful blockade solely by pinprick-induced analgesia in the perineal and anal regions, without evaluating motor blockade—a methodological divergence from Li et al.^[7]^, who used a composite endpoint of perineal cutaneous analgesia and anal sphincter relaxation. The primary rationale is that anal sphincter relaxation typically indicates motor nerve blockade by local anesthetics during caudal anesthesia. However, ropivacaine exhibits sensory-motor dissociation (i.e., preferential blockade of pain-transmitting Aδ/C fibers over motor Aβ fibers at low concentrations)^[30]^. Consequently, their findings do not represent the minimum analgesic concentration for caudal blockade. Since our study used pain disappearance as the efficacy endpoint to determine the minimum analgesic concentration for caudal blockade, the resulting EC50 was lower than that reported by Li et al. Thirdly, we employed ultrasound localization (rather than real-time guidance) for caudal blockade to accommodate our institution’s high-volume, fast-paced anorectal surgery workflow. While this approach reduces procedural time, it may marginally decrease success rates compared to real-time guidance, potentially introducing a confounding factor.

## 6. Conclusion

In summary, our findings demonstrate a significant sex-based difference in the EC50 of ropivacaine for ultrasound-guided caudal block in elderly patients, with female patients requiring a higher EC50 than their male counterparts. The future of perioperative medicine in the aging population remains challenging, necessitating more precise and tailored dosing strategies to optimize clinical outcomes in this demographic.

## Data Availability

All relevant data are within the manuscript.

## Acknowledgments

We would like to thank all nurses who worked in this study.

## Contributions

Liang Zhang and Hongyan Zhou contributed to study conception and design. Fu Wang and Yin He contributed to study conduct. Mingyu Qu and Wei Zhao contributed to data analysis. Liang Zhang, Hongyan Zhou and Fu Wang contributed to manuscript preparation.

## Funding

This work is supported by the Science and Technology Research Program of Chongqing Municipal Education Commission (Grant No. KJQN202515128), and Chongqing medical scientific research project (Joint project of Chongqing Health Commission and Science and Technology Bureau) (No. 2024MSXM134).

## Notes

### Competing Interest Statement

The authors have declared no competing interest.

### Clinical Trial

ChiCTR2200064111

### Funding Statement

Yes

### Author Declarations

Approved by the ethics committee of Chongqing Traditional Chinese Medicine Hospital (Approval No.: 2021-KY-65)

